# Evaluating the cost-effectiveness of new treatment strategies for the management of young infants with low- or moderate-mortality risk signs of possible serious bacterial infection: Framework and study methodology

**DOI:** 10.1101/2025.03.06.25323540

**Authors:** Charu C Garg, Yasir Bin Nisar, PSBI Cost-Effectiveness Study Group

## Abstract

**Background:** The World Health Organization (WHO) is coordinating two randomized controlled trials (RCTs) across three sites in Africa (Ethiopia, Nigeria and Tanzania) and four in Asia [Bangladesh, India (two) and Pakistan] to generate evidence on the optimal place of treatment for young infants with a single low-mortality risk sign of possible serious bacterial infection (PSBI) and switching antibiotic therapy from injectable to oral in young infants with moderate-mortality risk signs of PSBI. This paper presents the framework and methodology used to compare the costs and evaluate the cost-effectiveness of these strategies.

**Methods:** Cost analysis will be conducted from societal (both hospital and household) perspectives. Hospital direct medical costs (staff, medicines, consumables), direct non-medical costs (inpatient bed costs and transport expenses), and indirect operational costs (management, administration, non-consumables, training, and communication) will be gathered using hospital surveys. Household costs, including direct medical payments for treatment (registration, consultation, medications, consumables, and laboratory tests) and non-medical costs of transport, food and boarding, will be collected using household surveys. In both RCTs, combined hospital and household costs (for medical and non-medical) will be used to calculate the cost per sick young infant in each study arm. Effectiveness measures, based on the absence of adverse outcomes, will be used to determine incremental cost-effectiveness ratios. Household wage loss will estimate the household burden per treated child, and indirect hospital costs will highlight additional health system burdens.

**Discussion:** This cost-effectiveness framework evaluates PSBI treatment in young infants, integrating health system and household perspectives. It seeks to identify safe, economical regimes to reduce economic burdens, inform national budget impact, and potentially prompt WHO guideline revisions for better infant care in low-resource settings globally.

## Introduction

Around 2.3 million neonatal deaths occurred in 2022 worldwide, accounting for 47% of under-five deaths.^1^ In South Asia and sub-Saharan Africa, neonatal infections account for up to 37% of all neonatal deaths.^2^ The World Health Organization (WHO) recommends hospital referral for inpatient injectable therapy for at least seven days with supportive care, if needed, for young infants (0-59 days old) (YI) with any sign of possible serious bacterial infection (PSBI).^3 4^ However, in resource-limited settings, a large proportion of the families of YI with PSBI refuse hospital referral due to several barriers such as distance to the health facility, cost of hospitalization, daily wage loss during hospitalization period and cultural constraints resulting in newborn deaths.^5 6 7 8^

To address these barriers, several clinical trials^9 10 11 12^ and research have been conducted to assess the feasibility of implementation^13^ of the WHO PSBI guidelines^14^ in Africa and Asia to establish that YI with clinical severe infection (CSI) and pneumonia (subclassification of PSBI) treated with simplified outpatient antibiotic regimens are effective. Pooled analyses of three trials from Africa and Asia reported that a simplified treatment regimen using oral amoxicillin with gentamicin had better outcomes compared to other regimens in ambulatory settings.^15^ Secondary analysis of the African Neonatal Sepsis Trial (AFRINEST) further highlighted that infants with signs of CSI had a higher mortality rate when treated in a hospital compared to outpatient treatment and further divided these signs into low-or moderate signs based on mortality risk. ^16^

Cost-effectiveness analyses (CEA) have become vital in identifying affordable and effective interventions for the management of neonatal sepsis and reduction in neonatal mortality among vulnerable newborns. A study from Ethiopia assessed the costs of managing PSBI for YI at health posts when a referral was not feasible and found it cost-effective.^17^ Similarly, AFRINEST trial found management of PSBI with simplified antibiotic regimens cost-effective in the Democratic Republic of Congo, Kenya and Nigeria.^18^ A systematic review across 14 low-income countries estimated an average hospitalization cost for YI with sepsis (considering both provider and payer perspectives) at United States of America dollars (USD) 55.^19^ Another multi-centric implementation research in India estimated USD 18.5 as the health system cost for recommended and acceptable outpatient treatment of PSBI with simplified antibiotic regimens.^20^

This study presents a comprehensive cost-effectiveness methodology for two randomized controlled trials (RCTs) evaluating new treatment strategies for managing PSBI in young infants. The first trial^21^ will evaluate the appropriate place of treatment for YI with a single low-mortality-risk PSBI sign and the second trial^22^ will evaluate the duration of hospitalization for those with moderate-mortality-risk PSBI signs by switching from injectable to oral antibiotic therapy. Additionally, this framework calculates household costs across each study arm, offering a holistic perspective on the economic and clinical impacts of these interventions. The primary objective is to determine whether the intervention is superior or at least non-inferior to the control in preventing poor clinical outcomes of both RCTs.

## Methods

### Study design and participants

This study involves two individual, parallel arm open-label RCTs being conducted at multiple hospitals at seven sites in six countries - three sites in Africa (Ethiopia, Nigeria and Tanzania) and four sites in Asia [Bangladesh, India (two sites) and Pakistan]. ^21 22^ The study participants will be YI with low-or moderate-mortality-risk PSBI signs that seek treatment from selected hospitals in the study area and live within a catchment area where a follow-up up to 14 days can be accomplished.

#### Interventions and controls in the two RCTs

The RCT1 is evaluating whether experimental outpatient treatment (intervention) for YI with a single low-mortality-risk PSBI sign (low mortality risk signs: high body temperature ≥38°C, severe chest indrawing, fast breathing of ≥ 60 breaths per minute in 1-6 days old infants) with injectable gentamicin (once daily) for two days and oral amoxicillin (twice daily) for seven days can be as effective as the recommended care with injectable ampicillin (2-4 times daily), and injectable gentamicin (once daily) with supportive care for seven days, if needed (control). ^21^

The RCT2 is evaluating hospitalized YI with moderate-mortality-risk PSBI signs, alone or in combination [low body temperature (<35.5°C), movement only when stimulated, stopped feeding well) or multiple low-mortality risk PSBI signs. These infants are treated similarly for the first two days with injectable ampicillin (2-4 times daily, depending on the weight of the young infant) and injection gentamicin (once daily). At 48 hours after starting treatment, if they show (i) clinical improvement in terms of the absence of all signs of critical illness or CSI, and (ii) have a negative C-reactive protein laboratory test, they are randomized into two groups; one is discharged on oral amoxicillin (twice daily) for five days at home (intervention) and the second group continues injectable antibiotic therapy in the hospital for five more days (control).22

**Outcomes**: The primary outcomes for the two RCTs are defined below:

Poor clinical outcomes for RCT1 are ^21^

i. death at any time from randomization up to day 15 of initiation of therapy, or
ii. any sign of critical illness (no movement at all, unable to feed at all, or convulsions) or any sign suggestive of another serious infection, e.g., meningitis, bone or joint infection on day 2, 4 or day 8 post-randomization, or
iii. any new CSI sign on day 4 or day 8 post-randomization, or
iv. persistence of the presenting CSI sign on day 8 post-randomization.

Poor clinical outcomes for RCT 2 are^22^

i. death between randomization (day three of initiation of therapy) and day 15 of initiation of therapy, or
ii. presence of any sign of critical illness (no movement at all, unable to feed at all, or convulsions) or any sign suggestive of another serious infection, e.g. meningitis, bone or joint infection, on day four and eight of initiation of therapy, or
iii. presence of any sign of CSI on day eight of initiation of therapy

The effectiveness rates will be estimated for each treatment arm for both RCTs as follows: Proportion or percentage of the population that did not experience poor clinical outcomes.

Effectiveness Rate= (Positive Outcomes (O)/ Total Population (N))×100

Where the number of individuals with positive outcomes (O)=N−P. The total population (N) is the total number of individuals in each arm of the treatment group and the population with poor clinical outcomes (P) is the number of individuals who experienced poor clinical outcomes (e.g., death, deterioration, or other conditions described above).

### Framework for analysis

A common standardized protocol is being implemented across all study sites to ensure consistency in participant selection, interventions, and outcome assessments. The WHO coordinating team provides central training and oversight to harmonize activities, outcomes, and analytical procedures. All participating sites have extensive research experience, further supported by regular online and in-person meetings, data monitoring, and site visits by WHO monitors. Ethical approvals have been obtained from the WHO and institutional ethics committees. The study started on 24 June 2021 and data collection was completed on 19 August 2024. Written informed consent was taken at each site from households for cost surveys.

#### Costs framework

The cost will be assessed from both the health system and household perspective. From the health system perspective, the medical and non-medical costs of treatment will be determined at each study hospital across all sites in both intervention and control arms. Additionally, the cost from the household perspective will also be separated into medical and non-medical costs. The indirect administrative costs at the hospital and opportunity costs for households will also be estimated. The sample framework for costs at the hospital for each study arm in RCT1 and RCT2 is shown in Figures 1 and 2, respectively. These are based on implementation strategies defined in the protocol. ^21 22^ The costs are weighted by infant days under each trial and for control and intervention arms separately.

**Figure 1:**
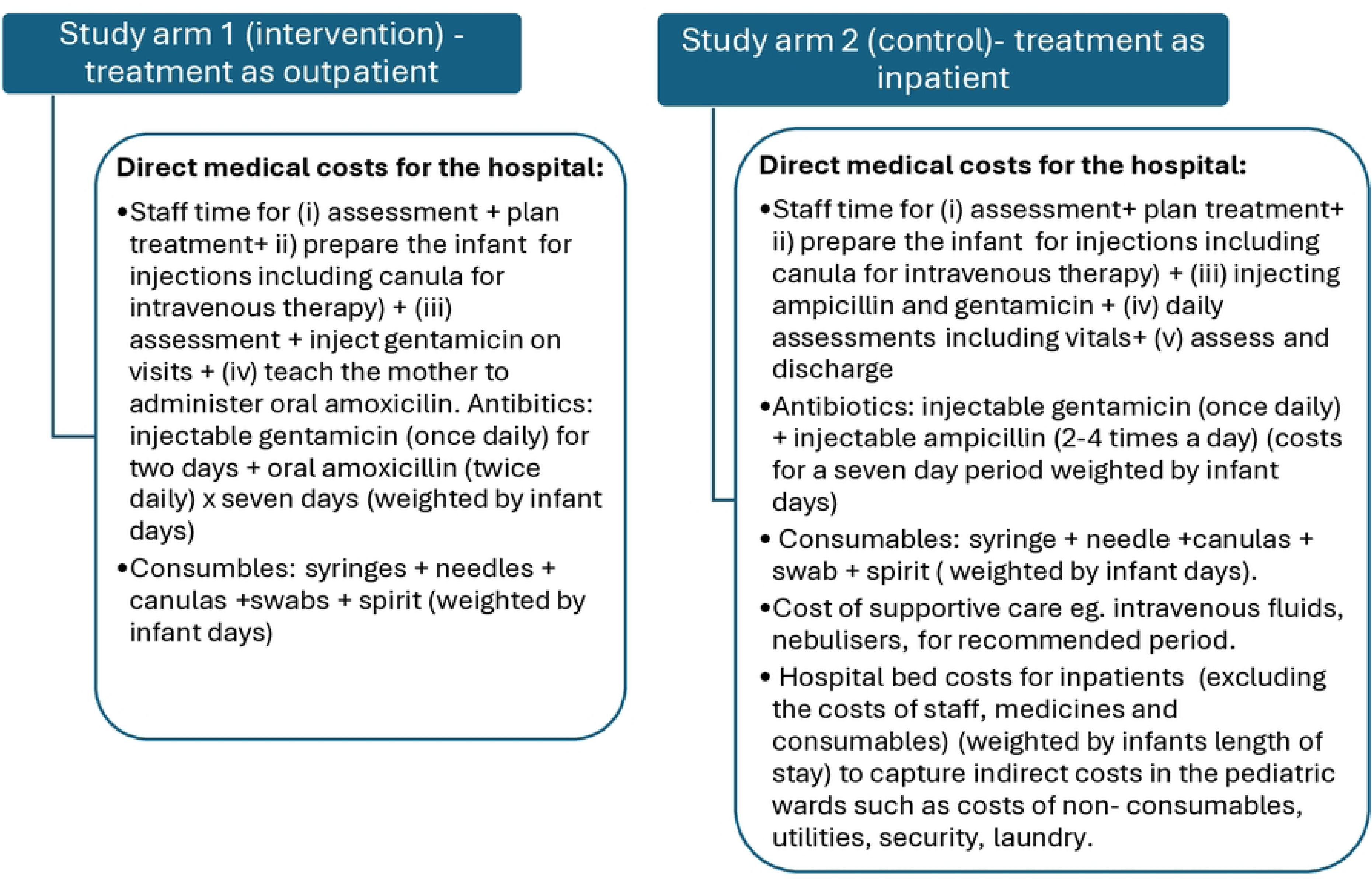
Sample framework for provider costs in each arm of RCT1

**Figure 2:**
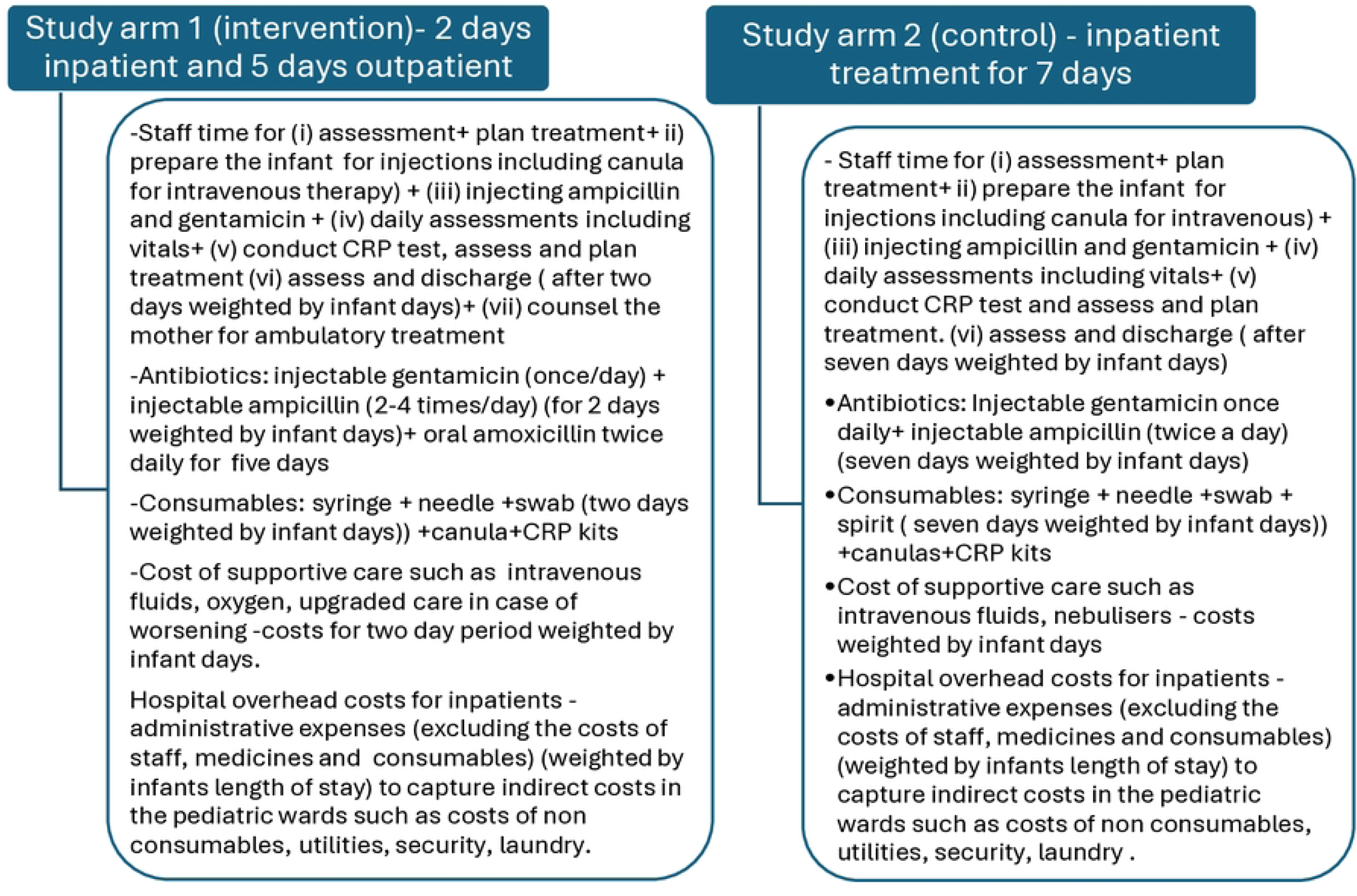
Sample framework for provider costs in each arm of RCT2

Figure 3 shows a framework for household costs for RCT1 and RCT2. The households incur out-of-pocket expenditures, which can be separated into direct medical expenses (registration/ consultation, medicines, consumables and laboratory) plus other non-medical expenses (transport, food, lodging). Additionally, there are costs for productivity lost in the form of wage loss. While direct medical and non-medical expenses will be used for CEA, costs for lost productivity will be reported separately. Weightage would be determined by the number of infant days spent on each expense item.

**Figure 3:**
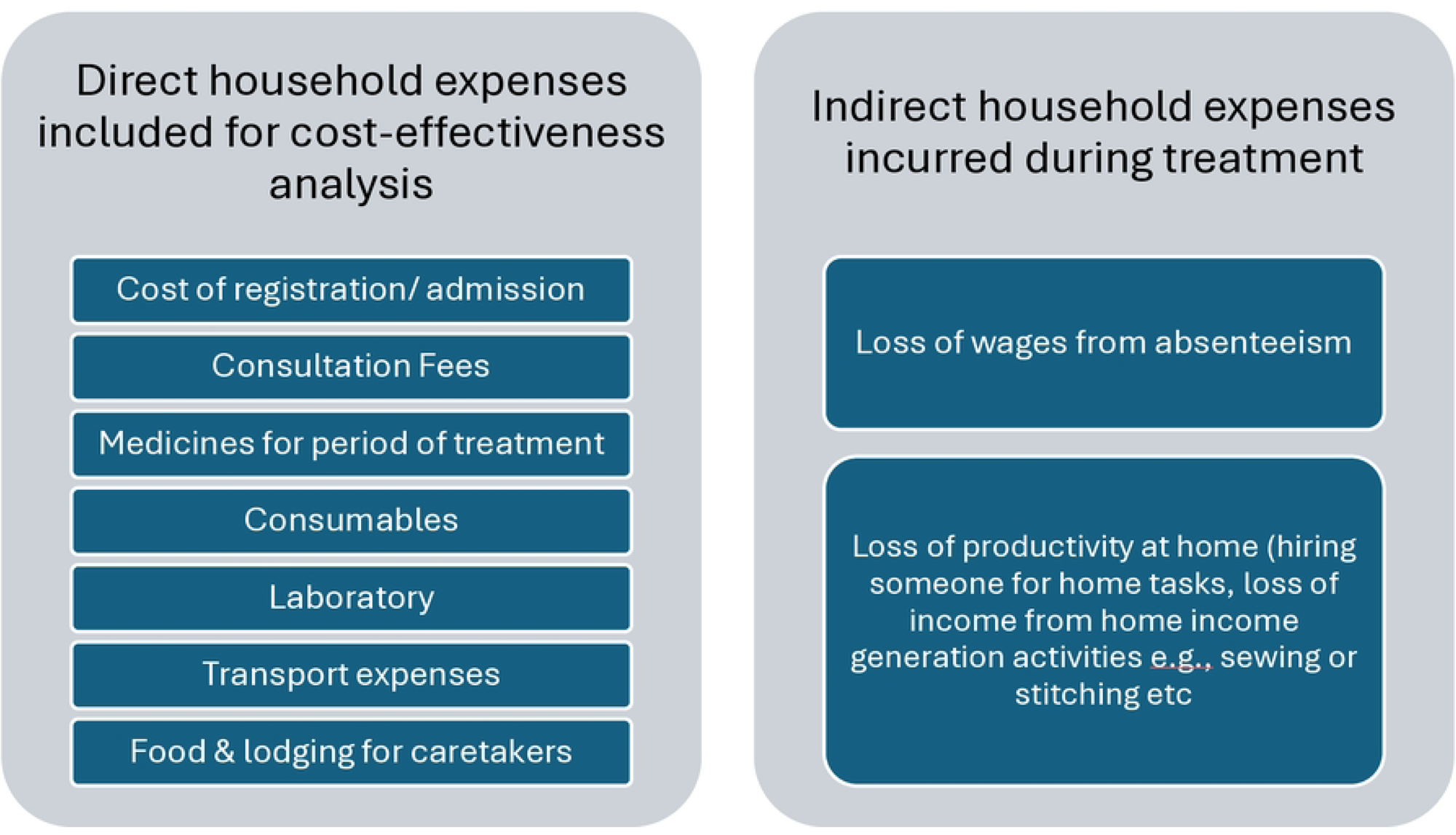
A framework with a comprehensive household perspective: out-of-pocket expenditures by households

### Data Collection

#### Hospital data

Provider data will be gathered using standardized hospital/provider questionnaires for both control and intervention arms in RCT1 and RCT2. Data will be collected from 28 hospitals from all seven sites (four hospitals in Bangladesh, four in Ethiopia, five in India Himachal Pradesh and National Capital region (India HP-NCR), six in India - Uttar Pradesh (India UP), three in Nigeria, four in Pakistan and two in Tanzania) where YI are enrolled for treatment under RCT 1 and RCT 2.

Provider data for CEA will include the following variables:

1. Coverage of sick young infants (SYI) assessed, enrolled and treated under both RCT 1 and RCT 2 for control and intervention.
2. Data on two outcome variables for all SYI covered under different treatment arms. A well-trained independent outcome assessor visits all enrolled cases on days 4, 8 and 15 after the initiation of treatment to assess the study outcomes in both intervention and control arms.
3. Average annual salaries of nurses, pediatricians, medical officers and any other staff supporting treatment. To calculate the staff costs per minute, data will be collected on average number of days in a month and average number of hours in a day that staff works.
4. Staff time for each staff type for each visit for an activity mentioned under the two frameworks (figures 1 and 2). The activities for the staff time will be based on the implementation strategy defined elsewhere.^21,22^ Staff time will be considered only for activities that are linked to hospital staff and not those linked to study staff such as outcome assessment or treatment documentation.
5. For each activity, data will be collected for the number of infants that receive the activity for at least 80% of the treatment - *recommended treatment*. For those that receive less than 80% of the treatment - *Partial treatment*, costs will not be calculated. The latter occurs due to a change in treatment, readmission or death. Those that undertake partial treatment may have a lower cost if they die or leave treatment early or may have a higher cost of treatment if shifted to another complicated treatment at the hospital.
6. Data on staff time per visit for an activity and the number of visits undertaken for each activity during the recommended treatment period will be collected by interviewing at least 3-5 staff of each type at each hospital.
7. The staff time for all visits during the treatment period will be weighted by the number of days an infant receives the activity and the number of YI receiving the activity (defined as weighted by infant days). For the first activity assessment on presentation at the hospital, the number of SYI will be all enrolled for a given treatment-whether partial or recommended. For all other activities, only those SYI who receive recommended treatment will be considered.
8. The average exchange rate used to convert the local currency costs into USD will be for the period the salaries are reported for providers.
9. The recommended treatment period will be taken as seven days for inpatient treatment in the control arm and seven days for outpatient treatment in the intervention arm for RCT1. For RCT 2, the average length of inpatient treatment is two days, prior to a negative C-reactive protein laboratory test. Thereafter for RCT2, the control will have 5 days of inpatient and intervention will be 5 days of ambulatory treatment. Sometimes the treatment in the hospital may extend to two and half days in the hospital before the SYI will be discharged to continue outpatient treatment.
10. The data on quantities used of injection gentamicin, injection ampicillin and oral amoxicillin will be as per recommended treatment per SYI for each treatment arm under each RCT.
11. The number of SYI receiving specific medicines for each arm under both RCTs will be assumed to be those receiving recommended treatment.
12. Information on types and quantities of consumables required with each medicine will be collected separately for each hospital and each arm under both RCTs.
13. The cost of medicines and consumables will be taken per size in which it is dispensed. Dosage for medicines varies with the weight of the YI. ^21,22^ Price will be taken for the full strength, and the rest will be marked as wastage. Price data will be taken from the hospital’s pharmaceutical department (if available), otherwise, the market price or WHO price will be used.
14. Data on annual expenditures and admissions in the department/s where YI are treated at the hospital will be collected to calculate the costs/ bed that can be allocated for utilities, laundry, security, data management, internet, non-consumables and other operational costs. Data will be collected for (i) total number of YI admitted during the year at the newborn intensive care units and pediatric wards for which the annual expenditure data is collected and (ii) the ratios of the average length of stay for all YI at the hospital and those SYI enrolled for RCT in order to allocate the total administrative costs. Expenditures will be calculated net of salaries, medicines and consumables to avoid double counting. The costs per admitted child is allocated to the enrolled SYI based on the ratio of the average length of stay.
15. Hospital ambulance to transport enrolled SYI under a specific treatment arm is a cost to the hospital. It is difficult to get this information separately for each enrolled SYI under different treatment arms from the hospital data. Indirect estimation based on the average cost of the trip will be used and the number of SYI using free public ambulance will be estimated from the household data for each treatment arm under RCT1 and RCT2.

Hospital data will also be collected for indirect health system/ operational costs which are not exclusively linked to the treatment of SYI for which CEA analysis will be conducted but help to strengthen the coverage and outcomes for the treatment of PSBI. These include

1. Salaries and time for the administrative staff who undertake activities, such as general management, supervision, coordination, logistics, social mobilization, monitoring and data management for the departments where all enrolled SYI are assessed and treated.
2. Staff training costs include the costs of development of training material and training cost per staff per day based on the per diems, honorarium for trainers, venue and refreshment costs. Total training costs will be calculated for the total staff that were used for treating the enrolled SYI, whether as outpatient or inpatient.
3. Communication costs including job aids or educational material, the cost of videos developed and played at the hospitals for educating the mothers about the danger signs.

While data will be collected on quantities of these items along with prices from the hospitals, these costs are not relevant for CEA and will be calculated only to understand the additional burden to the health systems.

#### Household cost data

Household expense data will be collected during the last 6 months of the study at each site, to capture the out-of-pocket expenditures that households incur during the treatment of enrolled SYI under both treatment arms for RCT1 and RCT2. Data will be collected from at least 10% of the SYI that are enrolled under each arm at each site. Standardized household questionnaires will be used across all sites to collect data on direct and indirect costs using the REDCap (Research electronic data capture) tool. ^23 24^ Training will be provided for collecting the data on these forms and pilots will help in improving the tool and data quality. Data inconsistencies will be identified, discussed with the sites and corrected at the analysis stage.

Data will be collected on the type of treatment arm and duration of treatment. Direct treatment costs incurred for each component shown in Figure 3 will be gathered in local currency from households that complete recommended treatment and incur expenses under each treatment arm for RCT1 and RCT2 at each site. Total direct medical expenses by households will be collected for registration and consultation fees, expenditures on medicines such as amoxicillin, ampicillin, gentamicin and other supportive care such as paracetamol; consumable supplies such as syringe, needle, canula, bandage, spirit, gloves; laboratory expenses such as x-ray, blood work, urine tests (investigation costs); or any other payments to hospitals. Non-medical expenses such as transport, food and lodging, will be recorded for 7 days. Data will include the mode of transportation and expenses incurred by any household member on round trips during the period of treatment. Similar information will be collected on food and boarding (near the hospital) with the amount spent outside the home during the days the infant had to stay at or visit the hospital.

Indirect expenses are opportunity costs, or the costs incurred by the household in terms of wage loss to take care of the sick child. Data will be collected from parents and caretakers who lost their wages due to time spent caring for the child. Household work that does not entail income will not be included as part of the opportunity cost. Data will be collected on type of occupation/source of income, average earnings during a day, month or year (in the ranges provided) and number of days not able to work to take care of the SYI.

### Statistical analysis procedures

The statistical analysis for the hospital cost data will be done in the Semper Curiosity TM platform – a cloud-based SaaS data capture and analytics tool.^25^ The data will be entered into the tool from hospital questionnaires. A specific program for the steps below will be used to calculate the average costs per SYI for staff, medicines, consumables, and inpatient bed costs. Mean, median and 95% confidence intervals will be calculated per SYI receiving recommended treatment for each of the cost variables at the hospital as well as at the household level. The hospital treatment costs will be weighted average of hospital cost variables where weights are infant-days or the number of enrolled SYI receiving recommended treatment under different components for the suggested number of days. Household direct medical costs plus transport food and lodging costs per SYI will be added to total hospital costs to estimate the total average costs for the control and intervention arms for RCT1 and RCT2 separately. The treatment outcomes between the control and interventions will be calculated for both RCTs to estimate incremental cost-effectiveness ratios.

#### Estimation of staff costs

The following steps will be followed to calculate the staff costs per SYI at a given site:

1. *Average Staff time per SYI* will be calculated first at each hospital for a given staff type for each activity by multiplying the time (t = minutes per visit) and the number of visits/ SYI for the activity during the treatment period (V). The number of visits for an activity per SYI can vary from one visit for assessment on admission to 21 visits for daily assessments, including vitals (generally three visits for 7 days of treatment). However, a given staff type, e.g. one nurse may take a longer time or do more or fewer visits for an activity than another nurse at the same or another hospital. Hence, the total time (T) per SYI for each activity (A) per provider type (P) at a site under different arms and trials will be

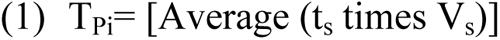

where ‘i’ is for providers of the same type to get the time and visits information, and ‘s’ is for each staff of a given provider type that is interviewed. Providers ‘i’ are nurses, medical officers/ general practitioners, pediatricians, or any other provider specified. We will take the average across the providers of the same type to get the average time for an activity during the entire treatment episode for each arm under both RCTs.

2. Staff time per provider ‘i’, per activity ‘j’ for a given hospital/facility (F) ‘k’ across all SYI getting recommended treatment will then be calculated as

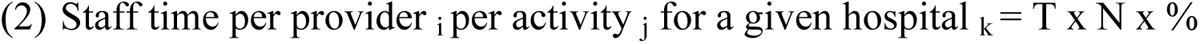

where N is the number of SYI receiving activity for full days of recommended treatment (N), and % is the percentage of infants treated by a given provider for the given activity. Activity ‘j’ is described in framework Figures 1 and 2 for RCT1 and RCT2, respectively, under control and interventions for staff time; ‘k’ is the hospital for which the data will be collected using the provider/hospital questionnaire.

3. The average time will be aggregated for all activities for a given provider and facility by using the number of SYI receiving recommended treatment as weights for each activity.
4. The total time for each staff will then be multiplied by respective staff salaries/ minute (calculated based on annual salaries, the number of days worked in a year, and the number of hours worked in a day) at each hospital to obtain the staff costs/provider for treating all infants completing recommended treatment at a hospital.
5. These staff costs will be added across the providers and then across the hospitals to obtain the total staff costs at a given site.
6. These staff costs will be divided by the total number of SYI treated under a given arm in each trial to get the staff costs/SYI for control and intervention separately for both RCT. These steps are pictorially represented in Figure 4 for staff cost calculations per SYI under control for RCT1. Likewise, staff costs per SYI will be calculated for each treatment arm under both RCTs.
7.

**Figure 4:**
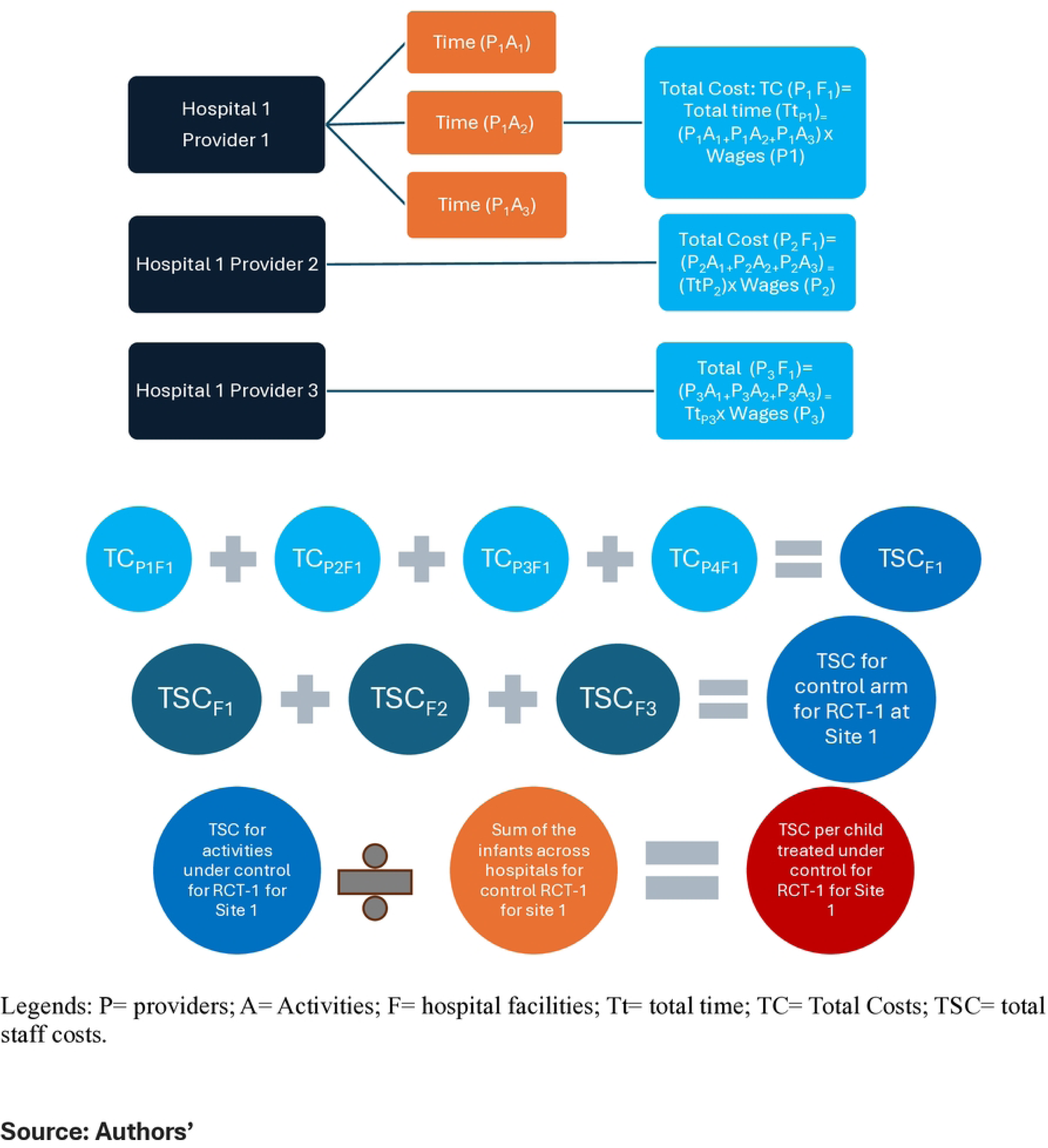
Pictorial representation for steps for calculating staff costs per sick young infant for a given arm for a given trial.

#### Estimation of medicines and consumables costs

The following steps will be followed to calculate the medicines and consumable costs per SYI at a given site:

1. For each medicine used under a given arm for a given trial, the total cost of a medicine for each SYI will be calculated by multiplying the price per vial or tablet for a medicine (P_d_) with the quantities of medicines required per SYI (Q_d_) for recommended treatment and number of infants receiving the medicine (n). Average medicine quantities will differ by age and weight of the SYI, or wastage with a given vial of injection or for tablets. For the hospitalized SYI, injection ampicillin is given twice daily in the first week of life, thrice daily in 2-4 weeks of life, and four times daily in four or more weeks of life. The dosage for each of the medicine is dependent on the weight of the infant. ^21 22^ For gentamicin, a vial of 40mg/ml or 20mg/2ml will be used per day, hence 7 vials will be needed for control arm and 2 vials for intervention arm under both RCT 1 and RCT 2. For ampicillin, we assume on an average 3 administration of 200 ml would be needed per day, hence an average of 2 ampicillin vials of 500 mg will be needed per day. For amoxicillin, 14 tablets of 250 mg will be required for 7 days treatment and 10 tablet for 5 days as outpatient for RCT 2.
2. For each medicine at each hospital, the cost of each consumable will be calculated by multiplying the price per unit of the consumable (P_c_) with consumable quantities required per SYI (Q_c_) for recommended treatment with that medicine, and the number of infants receiving the consumable (n).
3. The total cost of medicines will be estimated by adding the costs of all medicines at a given hospital needed for each arm for each RCT.
4. Similarly, the total cost of the consumables will be estimated by adding the costs of all consumables corresponding to the medicines needed for each of the arms under the two trials at a given hospital.
5. A few consumables are not linked to specific medicines but are linked to treatment of SYI, such as a cannula fixer, cannula for intravenous injections, gloves, CRP kits, etc. These costs will also be calculated by multiplying the price per unit of the consumable (P_c_) with consumable quantities required per SYI (Q_c_) for recommended treatment with that medicine, and the number of infants receiving the consumable (n) at the hospital.
6. The total cost of consumables will be estimated by adding the costs of all consumables at a given hospital needed for each arm under the two trials.
7. The total costs of medicines and consumables (estimated separately) at each site will be calculated by adding the costs at each hospital for each arm under the two trials.
8. Average weighted medicine and consumables costs at a site will be calculated by summing the costs of all medicines and all consumables (separately) across all hospitals for each treatment arm. These total costs of medicines and consumables (estimated separately) will be divided by the number of SYI at all hospitals at a site under each treatment arm to estimate medicine and consumable costs per SYI receiving recommended treatment at a hospital.

#### Estimation of the inpatient bed costs

1. Hospitals incur non-medical costs captured as inpatient bed costs. These are shared costs across all infants and children admitted to newborn intensive care units and pediatric wards for shared items such as non-consumables, utilities, laundry, security and any other expenses for these departments. The costs per admitted child is allocated to the enrolled SYI based on the average length of stay (ALOS).
2. First, the administrative expenditures per child admitted will be calculated by dividing the total annual administrative expenditures (exclusive of staff costs for treatment, medicines and consumables costs) of the pediatric and newborn intensive care departments by the total number of children admitted in the departments.
3. The ratio of the ALOS for the enrolled SYI to the total ALOS for children admitted to the hospital will be used as a weight to allocate the administrative expenditure per child to the enrolled SYI.
4. The inpatient bed costs per enrolled SYI will be estimated by multiplying the administrative expenditure/child estimated in step 1 with the weights calculated in step 2 for each hospital and for each arm under both trials. For the current analysis, we assume there are no inpatient bed costs for the intervention arm (outpatient) under RCT1 and for the 5 days ambulatory treatment in RCT2. Average weighted inpatient bed costs at a site will be estimated across all hospitals for each treatment arm by using the number of SYIs enrolled at the hospital as weights for a given treatment arm.

#### Estimation of transport costs for providers

1. Hospitals may incur ambulance costs for some patients brought to the hospitals. Since it is difficult to obtain data on the number of enrolled SYI that used public transport, an indirect estimation method will be used.
2. Household survey data will be used to determine the number of SYI who report using free public transport under different RCT treatment arms.
3. Each site will calculate the average cost per trip based on the average distance to the hospital and the cost per kilometer (km) or mile per trip. The cost per km is dependent on the salaries of the drivers, fuel for the average distance to the hospitals in the study area and the maintenance costs of the vehicle at the site.
4. Multiplying the average costs with the number of SYI under each arm will provide an estimate of transport costs.

#### Estimation of the household costs

1. Direct treatment costs reported by the households and the sum of the individual components will be compared, and data quality checks will be performed. A weighted average will be taken for the expenses incurred by the households on each of the components (registration, consultation, medications, consumables, and laboratory investigations) at the hospital/provider. Average costs incurred per SYI for each component will be calculated and reported.
2. Transport costs included as part of the CEA analysis will be calculated as average transport expenditures per SYI. Total expenses will be added for all days that transport is used by any caretaker for round-trip travel under each treatment arm and this is then divided by the number of households incurring expenditures whether for one day or more in that treatment arm.
3. For food and boarding charges, a similar method of estimation will be used as transport expenses. Total expenses reported by any caretaker during the treatment period will be added and then divided by the number of households incurring expenditures, whether for a day or more.
4. Opportunity cost of wage loss will not be used for CEA but reported separately. Wage loss will be presented (a) per family or per SYI (b) per member and (c) per day. The steps will be

a. Average wage/day will be calculated even when annual or monthly wages are given. The average wage for each caregiver will be the average of the range.
b. Average wage/day for each caregiver will be multiplied by the number of days lost for a SYI for any treatment. Maximum days lost by any caretaker will be capped at 15.
c. Sum of the total wage loss for a family under any treatment arm for all caretakers combined will be (A)
d. Sum up the number of SYI/ families that report wage loss for any of the caretakers will be
e. (B) under each treatment arm. SYI where no members will report a wage loss will be excluded from the denominator for calculations.
f. Sum of the total number of members that report a wage loss across different families will be (C).
g. Sum of the total number of days when a wage loss is reported will be (D).
h. Total wage loss per SYI or per family under each treatment arm= A/B
i. Total wage loss per member under each treatment arm= A/C
j. Total wage loss per day under each treatment arm= A/D

### Incremental Cost Effectiveness Ratios - standard methodology

Dividing incremental cost with incremental benefits will be used to calculate the incremental cost effectiveness ratio (ICER).

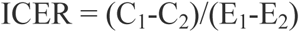

Where C_1_ are the average costs for the intervention group; C_2_ are the average costs for the control group; E_1_ is the outcome measure for the intervention group and E_2_ is the outcome measure for the control group.

There can be 4 case scenarios for ICER as shown in figure 5.

1. If C1<C2 and E1>E2, ICER is negative and falls in the southeast quadrant/zone of the CE plane in figure 5, and it implies that the intervention is more cost-effective than the control.
2. If C1>C2 and E1<E2, ICER is negative. This will fall in the northwest zone of the CE plane in figure 5 and conclude that control is more cost-effective.
3. If C1>C2 and E1>E2, ICER is positive. Here, intervention is more costly but more effective. This will fall in the northeast zone of the CE plane in figure 5. So, the decision about cost-effectiveness will depend on the cost-effectiveness threshold that varies by country.
4. If C1<C2 and E1<E2, ICER is positive, this will fall in the southwest zone of the CE plane in figure 5. Here, decisions usually remain inconclusive and, therefore, can be taken at the policy level if new interventions can be provided at a wider scale (owing to their low cost compared to existing treatment) to improve the coverage that will impact overall effectiveness in the population.

The ICERs with a negative value are relatively easy to interpret with values in the southeast quadrant implying new treatment is more effective and saves money compared with the old treatment. If ICER values are in the northwest quadrant, old treatment dominates the new treatment.

For each RCT, we will evaluate cost measures for control and interventions and compare each with effectiveness measures (non-poor clinical outcomes within 14 days). For each RCT, the ICER ratio will help to conclude if intervention is more cost effective as compared to control.

**Figure 5:**
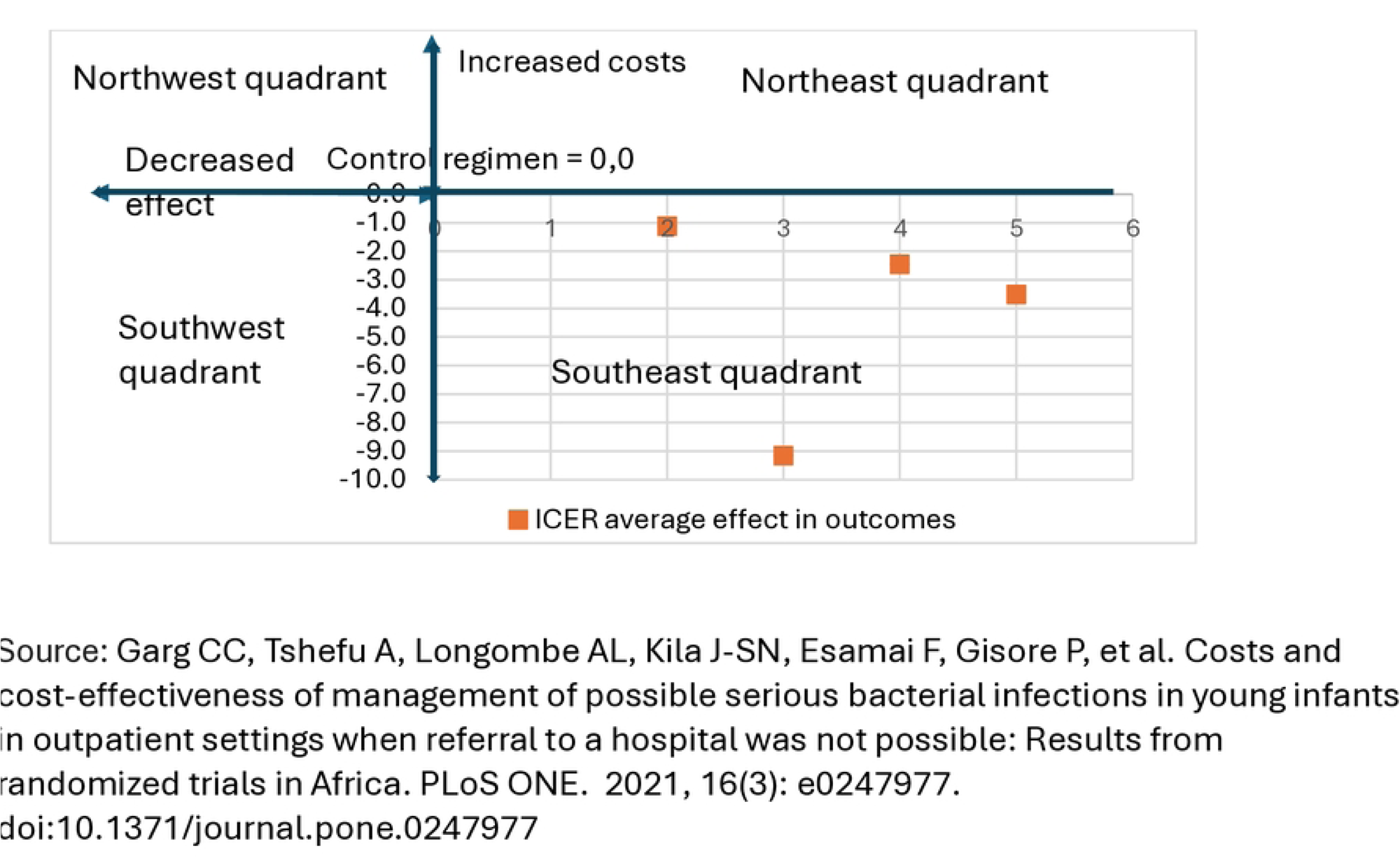
Incremental Cost effectiveness Ratio quadrants

## Discussion

This paper presents a detailed methodological framework for calculating the costs of managing PSBI in YI (SYI), integrating both health system and household perspectives across different contexts. It is the first study of its kind to comprehensively assess economic and clinical outcomes using such a dual approach. By analysing direct treatment costs (e.g., consultations, medicines, consumables) and direct non-medical costs such as transport, food, lodging, and indirect costs such as wage loss, this framework captures the full economic burden on households. Simultaneously, it evaluates hospital-level costs, including human and material resources, to estimate the economic strain on health systems. This methodology also facilitates comparisons of costs incurred for treating low-mortality-risk PSBI in both hospital and outpatient settings and for moderate-mortality-risk PSBI in SYI for a shorter and longer hospital stay. By establishing a standardized approach to CEA, the study enables more robust evaluations of RCT outcomes, supporting evidence-based decision-making and policy formulation. The findings from this study have the potential to reshape PSBI management guidelines.

For RCT1, if outpatient care for low-mortality-risk PSBI signs is shown to be safe and cost-effective, as compared to 7-day recommended inpatient treatment (control) it could significantly alleviate the burden on hospitals, allowing them to focus on critically ill young infants. This approach may also reduce the incidence of hospital-acquired infections while lowering costs for both health systems and households.

Similarly, RCT2 could demonstrate the feasibility of switching from the inpatient care of moderate-mortality-risk PSBI cases to outpatient care after a shorter hospital stay. If outcomes are shown to be non-inferior, this approach would optimize the use of hospital resources and minimize household economic burdens. Collectively, the results from these trials will provide a strong foundation for revising WHO guidelines, ensuring that treatment strategies align with resource-constrained settings while maintaining clinical safety.^14 26^

While this study provides a comprehensive assessment of costs from health systems and household perspectives, several limitations must be acknowledged. This analysis does not account for health system constraints, implementation barriers and capacity limitations or informal healthcare costs that families may incur. Additionally, key factors like supply chain inefficiencies, geographic cost variation, and quality of care metrics specific to PSBI treatment were beyond the scope of this framework. Addressing these limitations in future research will provide a more nuanced understanding of the costs and challenges associated with scaling these interventions.

This study’s findings have the potential to inform a paradigm shift in PSBI management. If outpatient care for low-mortality-risk PSBI signs and shorter hospital stays for moderate-mortality-risk cases are proven to be safe and cost-effective, these interventions could significantly reduce household and health system costs. Policymakers, providers, and payers would benefit from these insights, which could drive a revision of WHO’s PSBI treatment guidelines.^26^

## CONCLUSION

Current guidelines^26^ recommend seven days of hospitalization with injectable antibiotics for YI presenting with any sign of PSBI. However, in many infants in low- and middle-income countries (LMICs) access to care remains limited and adherence to referral is low due to financial, logistical or cultural barriers. Prolonged hospital stays impose significant economic burden on families and exacerbate resource constraints for already overburdened health systems in LMICs. By integrating both economic and clinical evidence, this study will contribute to the broader goal of improving neonatal survival and equity in healthcare access in resource-constrained settings.

## Data Availability

All relevant information are within the manuscript.

---

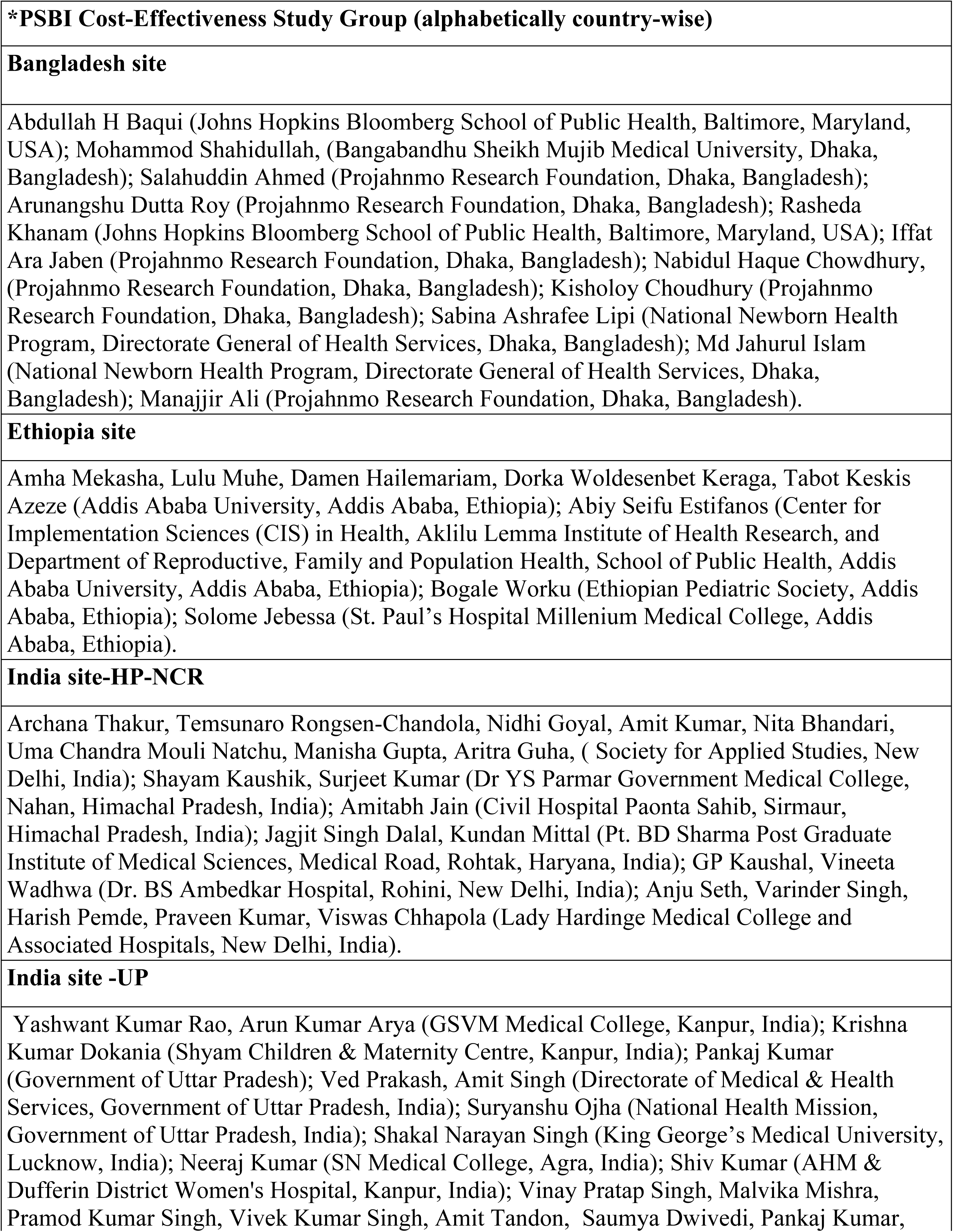
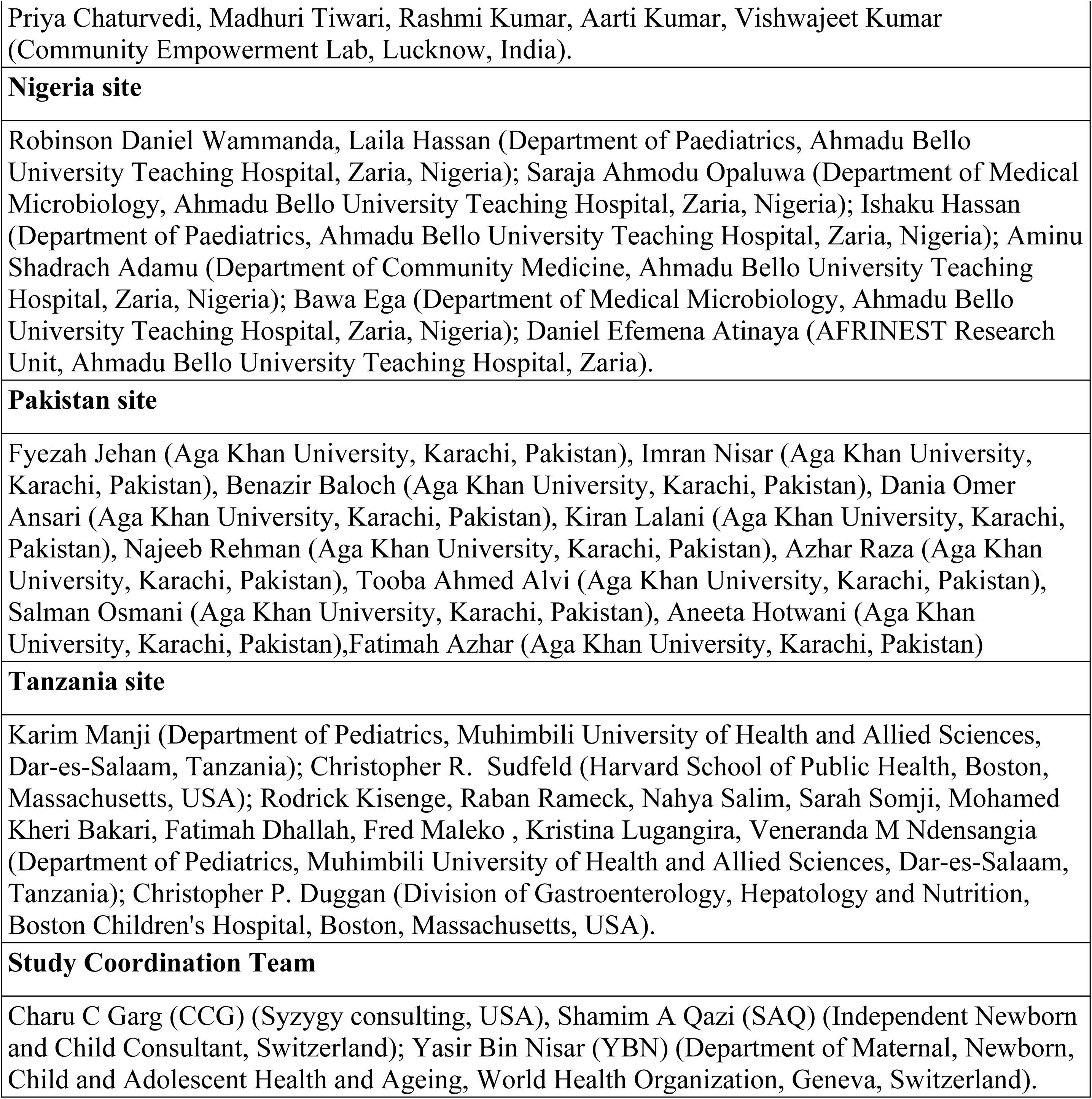

## Role of the PSBI study group: Contributors

CCG, YBN, and SAQ conceptualized and designed the cost-effectiveness study. All authors contributed to the interpretation of data and preparation of the manuscript. All authors approved the final version of the manuscript. More specific roles are in the table below:

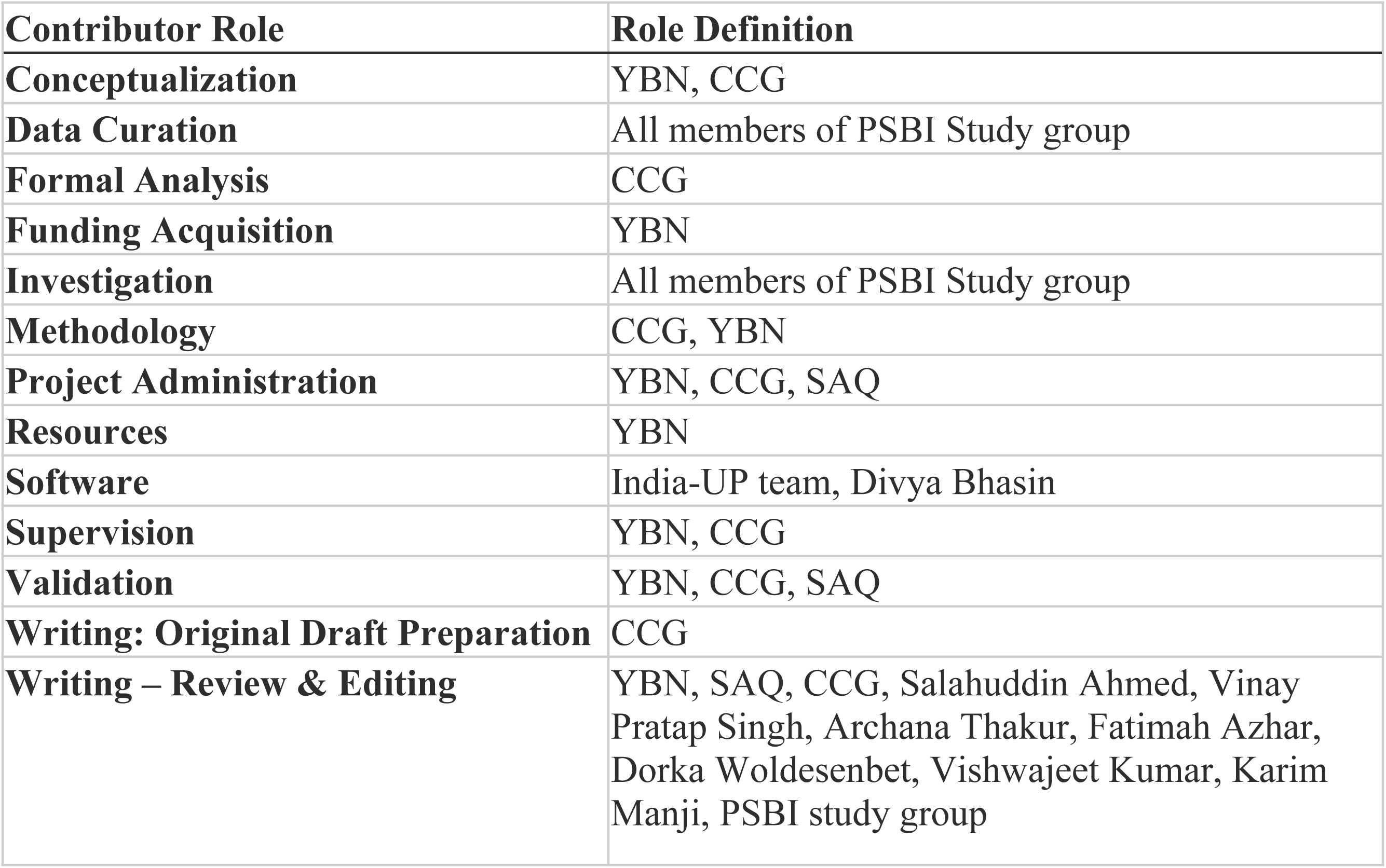

The study was funded by a grant from the Bill & Melinda Gates Foundation to WHO, which sponsored the study. We thank the families who participated in these studies, the project field staff for their hard work and dedication, and the physicians and administration at every site who provided data for the conduct of the study. The WHO appointed an independent data and safety monitoring board (DSMB) to monitor the quality of the data and protect the safety of infants enrolled in the trial. DSMB: William MacLeod (Chair), Harish Kumar Chellani, Simon Cousens, Haroon Saloojee and Fredrick Namenya Were.

Additionally, a Technical Advisory Group (TAG) was established by WHO. TAG: Jose C Martines (chair), Stephen Wall, Satinder Aneja, and Kristina Keitel. Divya D Bhasin (Cofounder, Semper Centrum) supported CCG to develop the methodology for hospital data analysis and Karina Gupta and Nikita Bindra (Research staff, Syzygy Consulting) supported with literature review and formatting.

## REFERENCES

1. United Nations Inter-agency Group for Child Mortality Estimation (UN IGME). Levels and trends in child mortality: Report 2023. New York: United Nations Children’s Fund, 2024 (Available from https://childmortality.org/wp-content/uploads/2024/03/UNIGME-2023-Child-Mortality-Report.pdf)

2. Alliance for Maternal and Newborn Health Improvement (AMANHI) Mortality Study Group Population-based rates, timing, and causes of maternal deaths, stillbirths, and neonatal deaths in South Asia and Sub-Saharan Africa: a multi-country prospective cohort study. Lancet Glob Health. 2018;6:e1297–308. [DOI] 10.1016/S2214-109X(18)30385-1 [PMC free article]

3. World Health Organization. Integrated management of childhood illness: management of the sick young infant aged up to 2 months. IMCI chart booklet. Geneva, Switzerland: WHO; 2019 Available from: https://www.who.int/maternal_child_adolescent/documents/management-sick-young-infant-0-2-months/en/. Accessed: 2024, November 5.

4. World Health Organization. Pocket book of hospital care for children: guidelines for the management of common childhood illnesses. 2nd ed. Switzerland: World Health Organization; 2013. Available from: https://apps.who.int/iris/bitstream/handle/10665/81170/9789241548373_eng.pdf?sequence=1. Accessed: 2024, November 5. [PubMed]

5. Leul A, Hailu T, Abraham L, Bayray A, Terefe W, Godefay H, et al. Innovative approach for potential scale-up to jump-start simplified management of sick young infants with possible serious bacterial infection when a referral is not feasible: Findings from implementation research. PLoS ONE 16(2): e0244192. 10.1371/journal.pone.0244192

6. Wammanda RD, Adamu SA, Joshua HD, Nisar YB, Qazi SA, Aboubaker S, et al. Implementation of the WHO guideline on treatment of young infants with signs of possible serious bacterial infection when hospital referral is not feasible in rural Zaria, Nigeria: Challenges and solutions. PLoS One. 2020;15:e0228718. 10.1371/journal.pone.0228718 [DOI] [PMC free article] [PubMed]

7. Rahman AE, Herrera S, Rubayet S, Banik G, Hasan R, Ahsan Z, et al. Managing possible serious bacterial infection of young infants where referral is not possible: Lessons from the early implementation experience in Kushtia District learning laboratory, Bangladesh. PLoS One. 2020;15:e0232675. 12. 10.1371/journal.pone.0232675 [DOI] [PMC free article] [PubMed]

8. Awasthi S, Kesarwani N, Verma RK, Agarwal GG, Tewari LS, Mishra RK, et al. Identification and management of young infants with possible serious bacterial infection where referral was not feasible in rural Lucknow district of Uttar Pradesh, India: An implementation research. PLoS One. 2020;15:e0234212. 10.1371/journal.pone.0234212 [DOI] [PMC free article] [PubMed]

9. Tshefu A, Lokangaka A, Ngaima S, Engmann C, Esamai F, Gisore P, et al. Simplified antibiotic regimens compared with injectable procaine benzylpenicillin plus gentamicin for treatment of neonates and young infants with clinical signs of possible serious bacterial infection when referral is not possible: a randomised, open-label, equivalence trial. Lancet. 2015;385(9979):1767–76.

10. Tshefu A, Lokangaka A, Ngaima S, Engmann C, Esamai F, Gisore P, et al. Oral amoxicillin compared with injectable procaine benzylpenicillin plus gentamicin for treatment of neonates and young infants with fast breathing when referral is not possible: a randomised, open-label, equivalence trial. Lancet. 2015;385(9979):1758–66.

11. Baqui AH, Saha SK, Ahmed AS, Shahidullah M, Quasem I, Roth DE, et al. Safety and efficacy of alternative antibiotic regimens compared with 7 day injectable procaine benzylpenicillin and gentamicin for outpatient treatment of neonates and young infants with clinical signs of severe infection when referral is not possible: a randomised, open-label, equivalence trial. Lancet Glob Health. 2015;3(5):e279–87.

12. Mir F, Nisar I, Tikmani SS, Baloch B, Shakoor S, Jehan F, et al. Simplified antibiotic regimens for treatment of clinical severe infection in the outpatient setting when referral is not possible for young infants in Pakistan (Simplified Antibiotic Therapy Trial [SATT]): a randomised, open-label, equivalence trial. Lancet Glob Health. 2017;5(2):e177–e85.

13. Nisar YB, Aboubaker S, Arifeen SE, Ariff S, Arora N, Awasthi S, et al. A multi-country implementation research initiative to jump-start scale-up of outpatient management of possible serious bacterial infections (PSBI) when a referral is not feasible: Summary findings and implications for programs. PLOS ONE. 2022;17(6):e0269524.

14. World Health Organization. Guideline: Managing possible serious bacterial infection in young infants when referral is not feasible. Geneva, Switzerland: WHO; 2015. Available: https://www.who.int/maternal_child_adolescent/documents/bacterial-infection-infants/en/. Accessed: 2024, November 5. [PubMed]

15. Longombe AL, Ayede AI, Marete I, Mir F, Ejembi CL, Shahidullah M, et al. Oral amoxicillin plus gentamicin regimens may be superior to the procaine-penicillin plus gentamicin regimens for treatment of young infants with possible serious bacterial infection when referral is not feasible: Pooled analysis from three trials in Africa and Asia. J Glob Health. 2022 Nov 21;12:04084. doi: 10.7189/jogh.12.04084. PMID: 36403158; PMCID: PMC9676044.

16. Nisar YB, Tshefu A, Longombe AL, Esamai F, Marete I, Ayede AI, et al. Clinical signs of possible serious infection and associated mortality among young infants presenting at first-level health facilities. PLOS ONE. 2021;16(6):e0253110.

17. Mathews B, Owen H, Sitrin D, Cousens S, Degefie T, Wall S et al. Community-based interventions for newborns in Ethiopia (COMBINE): Cost-effectiveness analysis. Health Policy Plan. 2017, 32, i21–i32. doi: 10.1093/heapol/czx054

18. Garg CC, Tshefu A, Longombe AL, Kila J-SN, Esamai F, Gisore P, et al. Costs and cost-effectiveness of management of possible serious bacterial infections in young infants in outpatient settings when referral to a hospital was not possible: Results from randomized trials in Africa. PLoS ONE. 2021, 16(3): e0247977. doi:10.1371/journal.pone.0247977.

19. Salman O, Procter SR, McGregor C, Paul P, Hutubessy R, Lawn JE, et al. Systematic review on the acute cost-of-illness of sepsis and meningitis in neonates and infants. Pediatr Infect Dis J. 2020;39(1):35–40. doi:10.1097/INF.0000000000002500.

20. Garg CC, Mukhopadhyay R, Arora NK, Awasthi S, Verma RK, Poluru R, et al. Cost of treating sick young infants (0-59 days) with possible serious bacterial infection in resource constrained outpatient primary care facilities: An insight from implementation research in two districts of Haryana and Uttar Pradesh (India). J Glob Health 2023;13:04062.

21. PSBI Study Group. Optimal place of treatment for young infants aged less than two months with any low-mortality-risk sign of possible serious bacterial infection: Study protocol for a randomised controlled trial from low-and middle-income countries. J Glob Health 2023;13:04055. Available from: https://jogh.org/2023/jogh-13-04055

22. PSBI Study Group. How long should young infants less than two months of age with moderate-mortality-risk signs of possible serious bacterial infection be hospitalised for? Study protocol for a randomised controlled trial from low-and middle-income countries. J Glob Health. 2023 Jul 14;13:04056. doi: 10.7189/jogh.13.04056. PMID: 37448340; PMCID: PMC10345886.

23. Harris PA, Taylor R, Thielke R, Payne J, Gonzalez N, Conde JG. Research electronic data capture (REDCap) – a metadata-driven methodology and workflow process for providing translational research informatics support. J Biomed Inform. 2009 Apr;42(2):377–81.

24. Harris PA, Taylor R, Minor BL, Elliott V, Fernandez M, O’Neal L, et al. The REDCap consortium: building an international community of software partners. J Biomed Inform. 2019 May *9*. doi:10.1016/j.jbi.2019.103208.

25. https://www.sempercentrum.com/our-products/

26. WHO recommendations for management of serious bacterial infections in infants aged 0–59 days. Geneva: World Health Organization; 2024. (available at: https://www.who.int/publications/i/item/9789240102903)

